# Alterations of lipid homeostasis in serum and white adipose tissue in morbid obese patients are partly reversed by the bariatric surgery

**DOI:** 10.1101/2023.06.12.23291122

**Authors:** Flore Sinturel, Simona Chera, Marie-Claude Brulhart-Meynet, Jonathan Paz Montoya, Etienne Lefai, François R. Jornayvaz, Giovanni D’Angelo, Minoa Karin Jung, Zoltan Pataky, Howard Riezman, Charna Dibner

**Affiliations:** Division of Thoracic and Endocrine Surgery, Department of Surgery, University Hospital of Geneva, Geneva, 1211, Switzerland; Department of Cell Physiology and Metabolism, Faculty of Medicine, University of Geneva, Geneva, 1211, Switzerland; Diabetes Center, Faculty of Medicine, University of Geneva, Geneva, 1211, Switzerland; Institute of Genetics and Genomics in Geneva (iGE3), Geneva, 1211, Switzerland; Department of Clinical Science, University of Bergen, Bergen, Norway; Proteomics Core Facility, EPFL, Lausanne, 1015, Switzerland; Institute of Bioengineering, School of Life Sciences, EPFL, Lausanne, Switzerland; INRA, Unité de Nutrition Humaine, Université Clermont Auvergne, Paris, France; Division of Endocrinology, Diabetes, Nutrition, and Therapeutic Patient Education, Unit of therapeutic patient education, WHO Collaborating Centre, Department of Medicine, University Hospital of Geneva, Geneva, 1211, Switzerland; Division of Visceral Surgery, Department of Surgery, University Hospital of Geneva, Geneva, 1211, Switzerland; Department of Biochemistry, Faculty of Science, NCCR Chemical Biology, University of Geneva, Geneva, 1211, Switzerland

## Abstract

Gastric Bypass surgery (GBS) represents a well-established approach to counteract human morbid obesity and its related comorbidities in modern countries. Beside its beneficial effect on weight loss and glucose homeostasis, emerging evidence suggests that GBS impacts on the circulating levels of phospho- and sphingolipids. However, long-term effects of GBS on lipid metabolism have not been explored. Thereby, we aimed to unveil to what extent GBS improves lipid homeostasis in serum and tissues from morbid obese individuals.

To investigate alterations in lipidomic signatures associated with massive weight loss following GBS in morbid obese patients, we employed direct infusion tandem mass spectrometry (MS) allowing to quantify a wide range of lipid metabolites in serum and subcutaneous adipose tissue (SAT) samples. Systematic lipidomic analyses were conducted in samples collected in a longitudinal cohort of patients (cohort 1, n = 11) prior to GBS, and one year following the surgery. These novel data were cross compared with our recent lipidomic analyses conducted by the same approach in an independent cohort of morbid obese patients and lean controls, where serum and visceral adipose tissue (VAT) lipids were analysed (cohort 2, n = 39).

Over 400 phospholipid and sphingolipid species have been quantified in serum and SAT (cohort 1), allowing to establish detailed lipidomic signatures associated with morbid obesity in a tissue-specific manner. Concomitant with weight loss and improvement of metabolic parameters, a massive rearrangement of lipid metabolites was observed one year following GBS. Strikingly, a substantial reduction of ceramide levels and increased amount of hexosylceramides were detected in both serum and SAT. The comparison of these new lipidomic profiles with the serum and VAT lipidomes established from lean and morbid obese subjects (cohort 2) revealed that GBS partly restored the lipid alterations associated with morbid obesity.

Our study provides the first systematic analysis of the long-term lipid homeostasis modifications upon GBS in humans SAT and serum and demonstrates that lipid metabolism alterations associated with morbid obesity might be partly reversed by GBS.

The research protocol was registered with the Protocol Registration and Results System at ClinicalTrial.gov [NCT03029572].

## Introduction

The Roux-en-Y gastric bypass surgery (GBS) is a commonly used treatment applied to counteract morbid obesity in modern countries. It usually leads to a drastic and fast reduction of weight and fat mass. Whereas weight loss and glucose metabolism improvement following GBS have been well-documented ^1-3^, long-term modifications of lipid metabolism following GBS stay largely unexplored. Indeed, all the studies conducted so far on the impact of the surgery-induced weight loss on the human plasma lipidome were performed 1-3 months following the surgery ^4-8^. Moreover, such metabolomic or lipidomic studies focused on a limited range of lipid metabolites analysed in the serum and did not examine the full spectrum of GBS-induced lipid changes. With the recent advances in lipidomics approaches, a broad range of individual lipid species can be at present quantified in human plasma and metabolic tissues, greatly expanding our knowledge regarding the complexity of lipid dysregulation in metabolic diseases ^9-13^.

We therefore established the systematic lipid signatures in a longitudinal cohort of morbidly obese subjects, conducted prior to, and one year after GBS (cohort 1). To this end, direct infusion tandem mass spectrometry-based lipidomic approach (MS) has been employed, allowing to quantify a wide spectrum of lipid metabolites. We here provide the first detailed analysis of the alterations of lipid homeostasis occurring in the subcutaneous adipose tissue (SAT) and the blood samples collected longitudinally from the same individuals prior to GBS and 1-year later. We completed these novel analyses by cross comparing the obtained data with our previous lipidomic analyses conducted by the same approach in a cohort of morbid obese patients in serum and visceral adipose tissue (VAT) samples (cohort 2).

## Results

### Alterations of clinical and metabolic features in morbid obese subjects one year following GBS as compared to the status prior to surgery (cohort 1)

Table supplement 1 reports the comparison of the main anthropometrical and serum profile characteristics of 11 morbid obese patients (cohort 1) before they underwent the surgery (pre-GBS), and approximately one-year following the surgery (post-GBS) ^14^. As previously described, the GBS induced a significant reduction of the total body weight, fat mass, waist, and hip circumferences, as compared to baseline morbid obese (pre-GBS) (Table supplement 1 ^14^). In addition, one-year post-GBS serum profiles exhibited a significant reduction of total blood cholesterol, LDL cholesterol (LDL-c), fasting insulin and leptin levels, as well as levels of several inflammatory biomarkers (Table supplement 1).

### Substantial alterations of the sphingolipid metabolism in serum following GBS (cohort 1)

Employing MS approach, we established detailed lipidomic signatures in serum samples collected before and one-year after GBS from the eleven patients (n = 22 paired samples). In parallel, five among these eleven patients were subjected to SAT biopsy collection before and one-year after GBS (n = 10 paired samples). Our lipidomic analyses detected a total of 400 lipid metabolites in sera and 413 in SAT samples across all the subjects. The lipidomics performed on the serum samples collected before and after the surgery from the same patients revealed 53 lipid metabolites statistically differentially abundant (FC > 1.5; p < 0.05) (Figure 1a). Hierarchical clustering performed on 100 top differentially abundant lipids (DALs), not all of which were qualified as significantly different, revealed a clear separation between the pre- and post-GBS samples (Figure 1b). Two classes of sphingolipids, Hexosylceramides (HexCer) and Ceramides (Cer), exhibited the most striking changes in the patient sera over a year-time post-GBS (Figures 1a-d). Decrease in Cer lipids was concomitant with an increase in the HexCer species after GBS (Figures 1d-f). Interestingly, the changes observed in mature sphingolipids (Dihydroceramides, DHCer) were not paralleled with a similar modification of the precursor sphingolipid forms (Hexosyldihydroceramides, HexDHCer) (Figure 1d), suggesting that the decrease in Cer levels may not stem from a downregulation of the *de novo* synthesis. The most striking decrease in Cer was observed for the most abundant species Cer40 and Cer42 (Figure 1e). In parallel, the level of the most abundant HexCer metabolite HexCer C42 was significantly increased in the post-GBS samples (Figure 1f). Although the total amount of SM was not modified after GBS (Figure 1c), the levels of the long-chain SM species were significantly reduced (Figure 1g). Overall, these results highlight a massive rearrangement of the sphingolipid metabolism during the one-year time following GBS.

**Figure 1.**
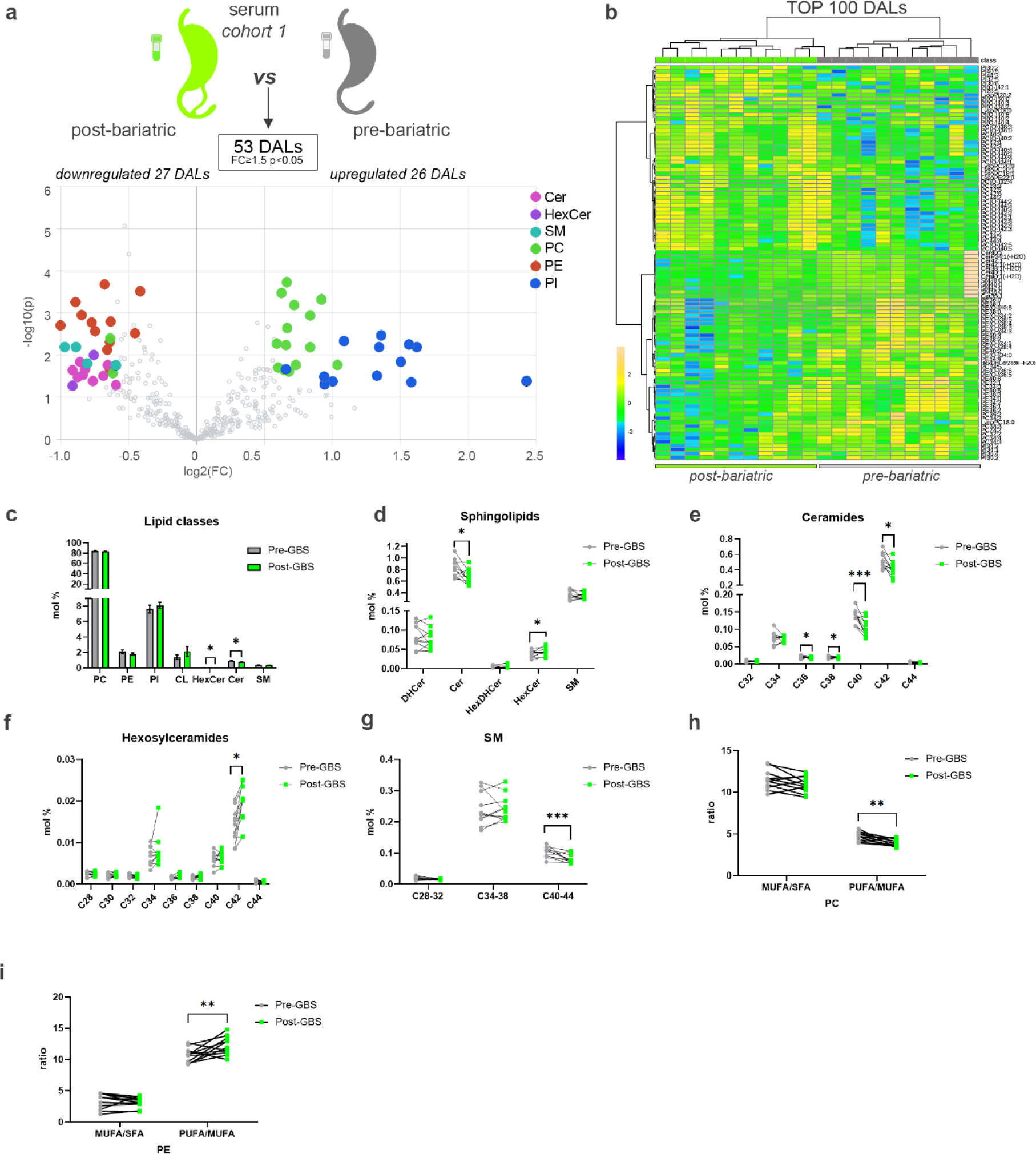
Comparison of differentially abundant serum lipids measured in morbid obese individuals before and after bariatric surgery (*cohort 1*) (**a**) Volcano plot of the DALs between pre- and post-bariatric serum samples (cohort 1, n = 11). Coloured dots highlight significant up- or down-regulated individual lipid species (fold change ≥ 1.5 and p < 0.05, paired design). (**b**) Hierarchical analysis *(Distance Measure: Euclidian; Clustering algorithm: Ward)* of top 100 serum lipids with most contrasting patterns between pre- and post-bariatric serum samples (n = 11). (**c**) Lipid class repartition (PC, PE, PI, CL, HexCer, Cer and SM) in human serum collected before and after GBS (in mol%). (**d**) Relative level changes (mol%) of DHCer, Cer, HexDHCer and HexCer in sera collected before and after GBS. (**e-f**) Relative Cer (**e**) and HexCer (**f**) level changes (mol%) in serum collected before and after GBS, represented according to the chain length. (**g**) Relative SM level changes (mol%) in sera collected before and after GBS, represented according to the chain length. **(h-i)** PC (**h**) and PE (**i**) ratios of MUFA to SFA, and of PUFA to MUFA detected in sera collected before and after GBS. Statistics for (**c-i**) are paired t-test. Data are represented as mean ± SEM. * p < 0.05; ** p < 0.01; ***p < 0.001.

Furthermore, both the hierarchical clustering and volcano plot analyses revealed significant changes in the levels of several serum Phosphatidylcholine (PC), Phosphatidylinositol (PI), and Phosphatidylethanolamine (PE) species following GBS (Figures 1a-b). Among the PC lipid species that exhibited altered levels post-GBS, we noticed an increase in Very Long Chain (VLC) PCs (Figure supplement 1a). Additionally, a decrease in the MUFA/PUFA PC ratio has been observed due to a significant decrease in PUFA species, along with an increase of SFA and MUFA PC species (Figure 1h and Figures supplement 1b-c). By contrast, the MUFA/PUFA ratio was increased, while the SFA decreased, within the PE group (Figure 1i and Figure supplement 1d). The ether PE species were significantly downregulated in the patient post-GBS sera samples as compared to their levels in pre-GBS counterpart (Figure supplement 1e).

### In human SAT, PI and HexCer metabolite levels are increased following GBS (cohort 1)

In the SAT samples, overall changes in the measured lipid levels were milder than those observed in the sera from the same individuals (Figures 2a-b). Indeed, the total amount of lipids clustered by lipid class did not significantly differ (Figure 2c). Volcano plot analysis revealed that 17 individual lipids were significantly altered, and the top 40 DALs showed a distinct separation between the sample groups (Figures 2a-b). The upregulated lipid metabolites were mainly represented by PI and sphingolipids, in particular by the HexCer (Figures 2a-b). Indeed, we observed an increase in these lipid species, as well as in HexDHCer, although the differences in their total amount did not reach statistical significance (Figure 2d). The total amount of the Cer was unchanged in n = 2 individuals, whereas it was decreased in 3 others post-GBS (Figure 2d). By contrast, 3 DHCer species (DHCer42:1(-H_2_0) DHCer42:0(-H_2_O) and DHCer40:0(-H_2_0) were significantly more abundant in post-GBS SAT samples as compared to pre-GBS counterpart (Figures 2a-b). Several Long-Chain (LC) SM species were increased as well, resulting in an opposite trend to the one that was observed in the serum (Figure supplement 2a).

**Figure 2.**
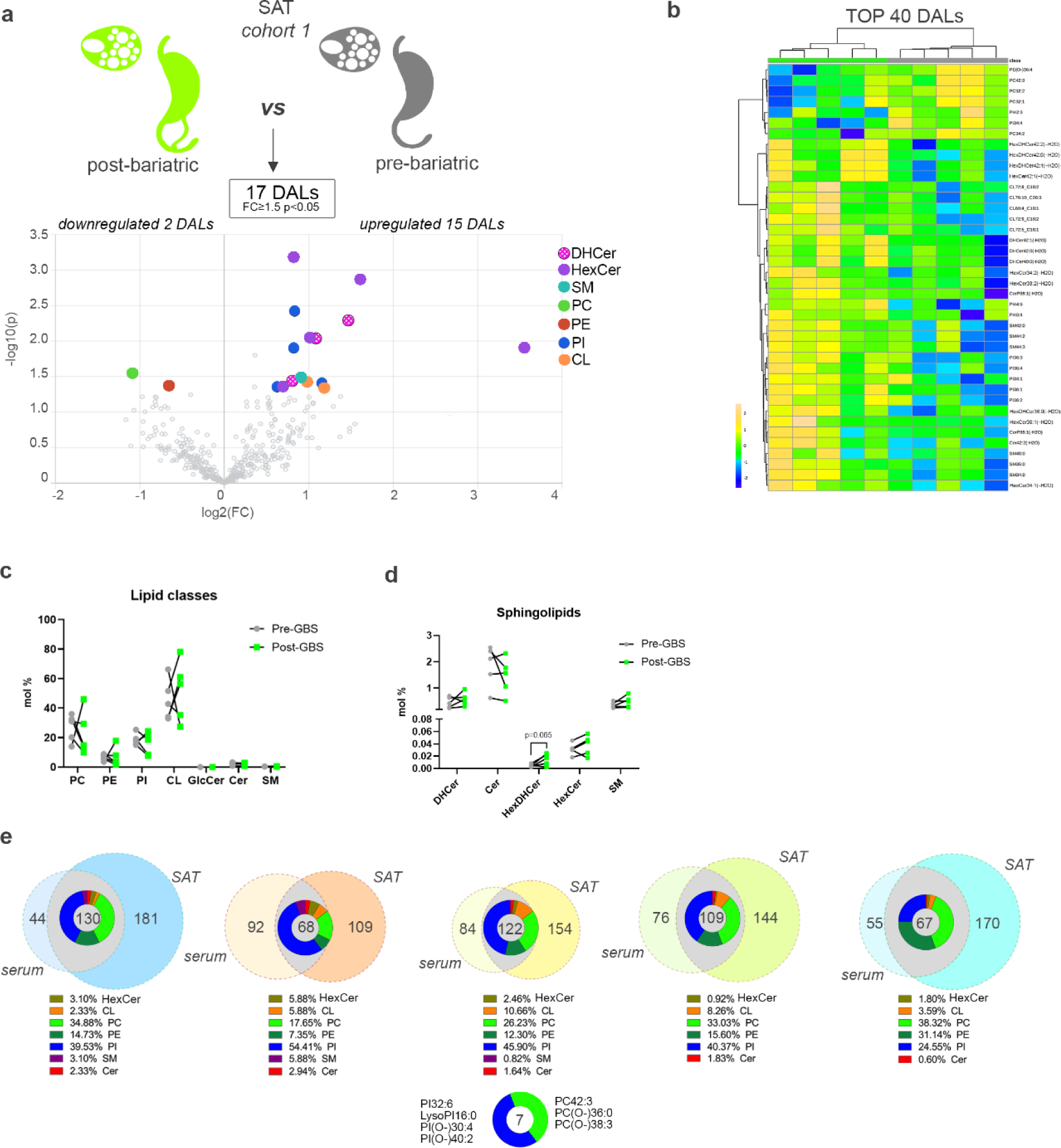
Comparison of differentially abundant SAT lipids measured in morbid obese individuals before and after bariatric surgery (*cohort 1*) **(a)** Volcano plot of the differentially abundant lipids between pre- and post-bariatric SAT samples (n = 5). Coloured dots highlight significant up- or down-regulated individual lipid species (fold change ≥ 1.5 and p < 0.05, paired design). **(b)** Hierarchical analysis *(Distance Measure: Euclidian; Clustering algorithm: Ward)* of top 40 SAT lipids with most contrasting patterns between pre- and post-bariatric SAT samples (n = 5). **(c)** Lipid class repartition (PC, PE, PI, CL, HexCer, Cer and SM) in human SAT collected before and after GBS (in mol%). **(d)** Relative level changes (mol%) of DHCer, Cer, HexDHCer and HexCer in human SAT collected before and after GBS. **(e)** Venn diagrams assessing the overlap between SAT and serum DALs (fold change ≥ 1.5 and p < 0.05) in each subject following GBS. Pie diagrams depict the range of lipid classes regulated in both SAT and serum following GBS. Seven lipids were differently regulated in both SAT and serum across all 5 analysed individuals. Statistics for (**c-d**) are paired t-test. Data are represented as mean ± SEM.

### The levels of PI metabolites are increased one year following GBS in both serum and SAT of morbid obese patients (cohort 1)

Comparison between the lipid composition of serum and SAT revealed a clear difference in the lipid class repartition. Indeed, CLs were predominantly observed in SAT, whereas PC lipids were mainly found in serum (Figure 1c and Figure 2c). In spite of such tissue specificity of lipid composition, a substantial overlap between DALs was observed in both serum and SAT for all the five subjects where we could collect both tissues (Figure 2e). In addition, seven lipids (3 PC and 4 PI metabolites) were commonly differentially regulated across all donors and all tissues (Figure 2e). Moreover, while the PI constituted only 8 to 20% of all the lipids detected in both serum and SAT (Fig 1c and Figure 2c), they represented, on average, more than 40% of the serum and SAT overlapping DALs (Figure 2e), revealing that PI lipid metabolites overcame the most pronounced alterations in both serum and SAT following GBS.

### Massive alteration of the serum and visceral adipose tissue (VAT) lipid landscape upon morbid obesity (cohort 2)

In order to complete our investigation of the GBS effects on the lipidomic features of morbid obese subjects, we re-analysed the serum and VAT lipidomes conducted recently ^9^, in a sub-cohort of lean and morbid obese individuals (cohort 2) (Figures 3 and 4, Table supplement 2). In sera, a number of individual PC lipids exhibited decreased levels in morbid obese individuals compared to the control samples (Figure 3a), although no significant difference was observed for the overall PC lipid class alterations (Figure 3b). Strikingly, while the long chain (LC) PC metabolite levels were significantly decreased in serum of morbid obese patients, in the levels of very long chain (VLC) PCs were significantly increased (Figure 3c). Similarly, we observed a diminution of the PUFA 2 PC lipids in morbid obesity, paralleled with upregulation of PUFA 4 (Figure 3d). Additionally, the sera obtained from obese individuals contained increased SM lipid levels (Figures 3a-b), in particular C34-38 SM lipids (Figure 3e).

**Figure 3.**
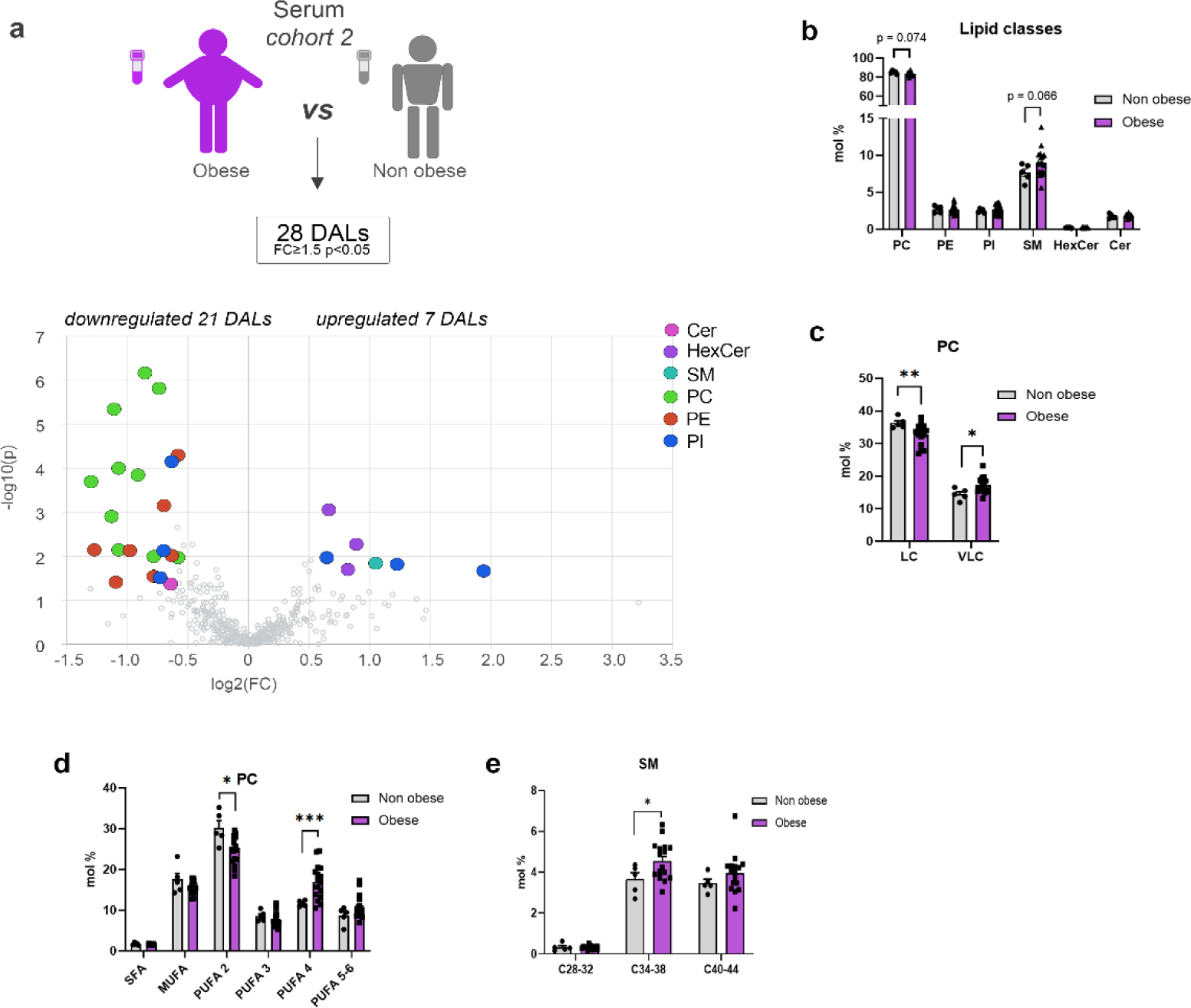
Serum lipids measured in morbid obese and lean control individuals (*cohort 2*) **(a)** Volcano plot of the differentially abundant lipids in serum between obese and lean control subjects. Coloured dots highlight significant up- or down-regulated individual lipid species (fold change ≥ 1.5 and p < 0.05, Welch’s corrected). **(b)** Lipid class repartition (PC, PE, PI, CL, HexCer, Cer and SM) in human serum from lean control and obese individuals (in mol%). **(c)** Relative PC level changes (mol%) in serum collected from lean control and obese individuals, clustered according to the chain length, LC (long chain C28-34), VLC (very long chain C38-44). **(d)** Relative PC level changes (mol%) in serum collected from lean control and obese individuals, represented according to the degree of saturation. **(e)** Relative SM level changes (mol%) in serum collected from lean control and obese individuals, clustered according to the chain length. Statistics for (**b-e**) are unpaired student’s *t*-test *P*-values. Data from obese (n = 16) and control (n = 5) individuals are represented as mean ± SEM. * p < 0.05, ** p < 0.01; ***p < 0.001.

**Figure 4.**
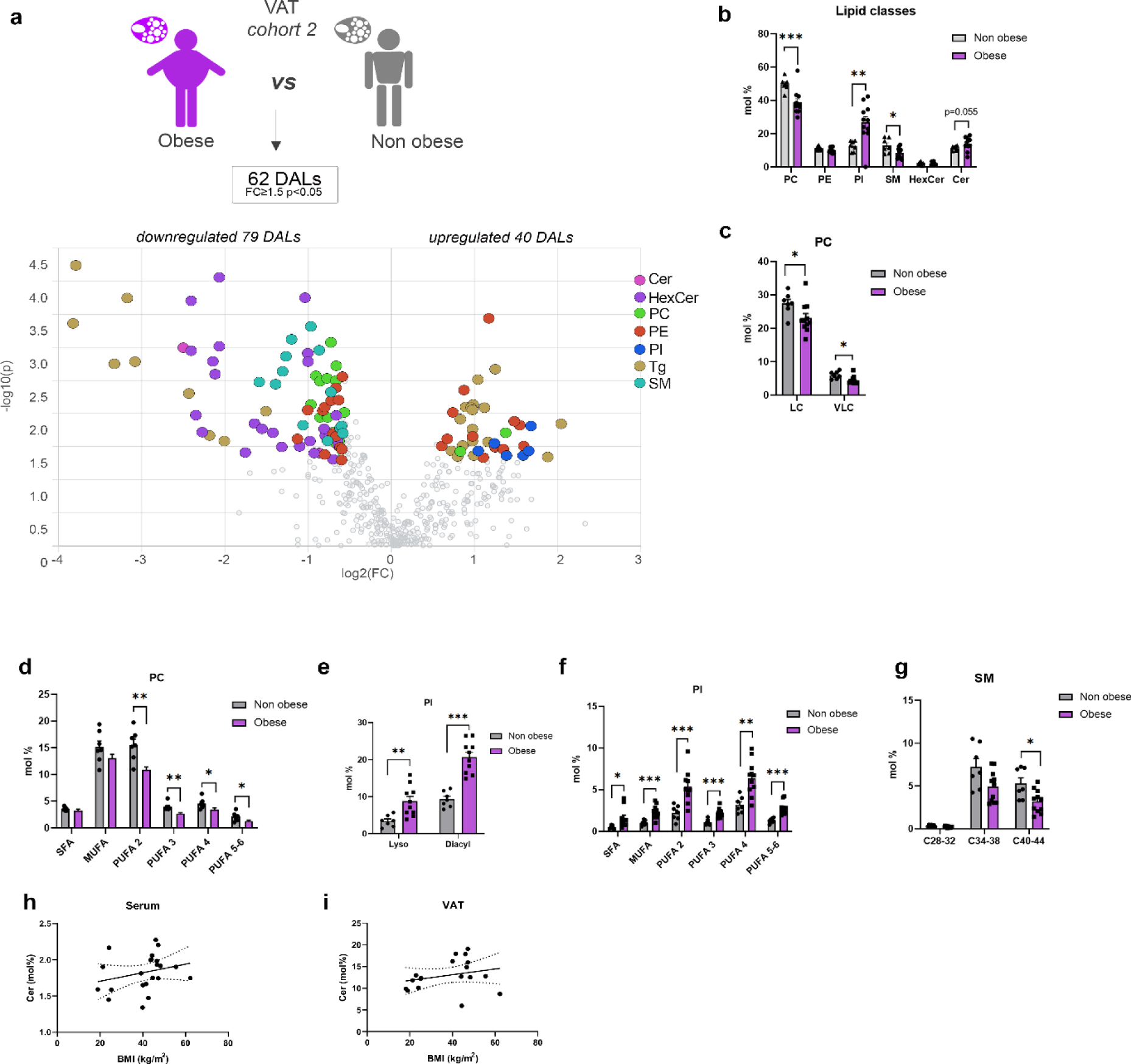
VAT lipids measured in morbid obese and lean control individuals (*cohort 2*) **(a)** Volcano plot of the differentially abundant lipids in VAT between obese and lean control subjects. In addition of the lipid classes measured in the other samples, Triglycerides (Tg) were measured. Coloured dots highlight significant up- or down-regulated individual lipid species (fold change ≥ 1.5 and p < 0.05, Welch’s corrected). **(b)** Lipid class repartition (PC, PE, PI, CL, HexCer, Cer and SM) in human VAT from lean control and obese individuals (in mol%). **(c-d)** Relative PC level changes (mol%) in VAT collected from lean control and obese individuals, clustered according to the chain length (**c**), and represented according to the degree of saturation (**d**). LC (long Chain C28-34), VLC (Very Long Chain C38-44). **(e-f)** Relative PI level changes (mol%) in VAT, represented according to the nature of the fatty acid linkage (diacyl or monoacyl (lyso)) (**e**), and to the degree of saturation (**f**). **(g)** Relative SM level changes (mol%) in VAT collected from lean control and obese individuals, clustered according to the chain length. **(h-i)** Association between the relative levels of ceramides detected in sera (**h**) or VAT **(i**), and the BMI of the subjects (serum n = 21, VAT n =17, Spearman correlation, for the serum R = 0.412, *p* = 0.057, for the VAT R = 0.315, *p* = 0.2). Statistics for (**b-g**) are unpaired student’s *t*-test *P*-values. Data from obese (n = 11) and control (n = 7) individuals are represented as mean ± SEM. * p < 0.05, ** p < 0.01; ***p < 0.001.

Noteworthy, the most striking alterations in the lipid metabolism upon morbid obesity were observed in VAT. Indeed, morbid obese individuals exhibited decreased levels of PC lipids (Figures 4a-d), similarly to the serum, associated with increased levels of PI metabolites compared to their control counterparts (Figures 4a-b, e-f). All the PC lipids were decreased, irrespective of the chain length or the degree of saturation of the species (Figures 4c-d). In contrast, the lyso- and diacyl PIs at all levels of saturation were overrepresented in VAT of morbid obese individuals (Figures 4e-f). In addition, there is an unambiguous diminution of SM lipids and of many HexCer metabolites in obese individuals compared to their control counterparts (Figures 4a-b, g). By contrast, Cer levels were elevated in the VAT of morbid obese individuals (Figure 4b).

### Bariatric surgery partly restores the lipid metabolism imbalance observed upon morbid obesity

A comparison between the lipid profiles observed in the sera and VAT of the cohort 2 subjects (Figures 3-4), and those obtained in pre- and post-GBS patient samples from the cohort 1 (Figures 1-2) suggests that the GBS partially rescues the lipidome landscape changes associated with obesity. Indeed, we observed a clear inversion of the sphingolipid phenotype associated with obesity, which was characterized by lower levels of SM and HexCer lipids (Figures 4a-b, g), in the patients following GBS (Figures 2a-d). Although the difference in Cer levels in VAT derived from obese subjects compared to the control counterpart did not reach statistical significance (Figure 4b), we detected a trend for an association between the patient BMI and the percentage of Cer measured in serum and VAT (Figures 4h-i). To a lesser extent, this correlation was conserved in our cohort of obese individuals who underwent GBS (Figure supplement 2b), while the levels of HexCer were inversely associated with the BMI (Figure supplement 2c). This finding suggests that the accumulation of Cer during the weight gain can be reversed by GBS, possibly via a conversion of Cer into HexCer. Concerning the glycerophospholipids, no clear inversion of the obese phenotype upon GBS has been observed. Indeed, high levels of VLC PC were detected in sera from morbid obese subjects (cohort 2, Figure 3c), and morbid obese patients one year after GBS (cohort 1, Figure supplement 1a). Similarly, the weight loss induced by the surgery was not sufficient to counteract the PI accumulation in SAT that characterized morbidly obese subjects (cohort 2, Figure 4b), with the high abundance of some PI species still observed one year following GBS (cohort 1, Figures 1a-b, and 2a-b).

## Discussion

Our study provides the first in-depth lipidome profiling of dynamic changes of the lipid homeostasis in serum and SAT from morbid obese patients one-year following GBS. The spectrum of DALs clearly differed between pre- and post-GBS samples, and was not identical in serum and SAT, highlighting the lipid tissue-specific changes upon GBS. Furthermore, the comparison of the lipidomic profiles from the morbid obese patients who overcame GBS (cohort 1) with our recent lipidomic analyses conducted in an independent cohort of morbid obese and lean control individuals (cohort 2), suggests that GBS partially rescues the alterations of lipid homeostasis associated with morbid obesity in humans (Table supplement 3).

So far, most of the studies reporting lipid changes upon GBS were conducted only in serum samples, and used targeted detection methods that cover a limited range of “classic” lipid species ^5-7, 15^. Here, we present the first in-depth characterization of the lipid profile of the serum and SAT of morbid obese human patients who overcame the GBS. Importantly, in contrast to previous studies conducted soon after GBS ^5, 6, 8^, we aimed at analysing sustained lipid metabolite changes over a one-year period following the surgery. Thus, the resulting lipidomic profiles reflect not only the impact of the acute weight loss effect, but also the long-term metabolic adaptations that were associated to this change in serum and in SAT. Our data support some of the previously proposed serum lipids as hallmark of weight loss following GBS. Thus, we confirmed a substantial decrease in serum Cer ^5, 6, 8, 15^ and LC SM ^4, 6^ species upon GBS (Figures 1c-e, g). Strikingly, our data demonstrate, for the first time, that the substantial alterations in the lipid landscape in morbid obese patients following GBS comprise the increased levels of HexCer (Figures 1d, f). Altogether, our findings indicate that a shift of the sphingolipid metabolism from Cer towards HexCer may take place during one-year period following GBS. Concerning the phospholipids, our results contrast with some recent works focused on serum lipid metabolic changes early after GBS. Indeed, although we consistently reported a significant decrease of ether PE in the serum one-year following the surgery (Figure supplement 1e) ^8^, we did not observe anymore a widespread decrease in PCs and/or PIs ^4, 6, 8^ in this context. This suggests that the diminution in these lipid classes is primarily associated with acute weight loss rather than reflecting a long-term effect.

One strength of the present study is the comparison of the lipidomic data obtained from samples collected upon GBS, with our recent lipidomic analyses of the lipid alterations associated with morbid obesity in performed in an independent human cohort ^9^. In the sera of morbid obese individuals, the levels of two choline containing phospholipid classes were drastically changed: an overall decrease in PCs was concomitant with an increase in SMs (Figures 3a-e). This observation agrees with previous studies showing that SMs accumulate in the serum of patients suffering from severe obesity and insulin resistance ^16-18^. Similarly, the levels of several PC species in the serum were recently found to be inversely associated with obesity ^10^. Strikingly, these obesity-related changes in the lipid metabolites were partly reversed in the patients one year following GBS (Table supplement 3). Of note, as SMs represent major components of LDL and VLDL particles ^19^, the lower abundance of serum SMs species post-GBS may reflect the significant reduction of plasma concentrations of LDL (Table supplement 1). Additional substantial change observed in post-GBS samples was the reduction of Cer levels, concomitant with increased HexCer amounts in serum, and to a lesser extent in SAT (Figures 1a-f, Figures 2a-d). Cer are well-known to accumulate during obesity, with decreased Cer levels being associated with increase in insulin sensitivity in mice and humans ^20-23^. In this line, we hereby report that Cer lipids are more abundant in the VAT of morbid obese individuals as compared to lean counterparts (Figures 4a-b). By contrast, we observed a significant upregulation of 3 DHCer species in SAT after the surgery (Figures 2a-b). The MS analyses that we employed here cannot distinguish DHCer lipid species from the toxic Deoxyceramide (DeoxCer) metabolites that were recently identified in the human adipose tissue ^9, 11^. Indeed, DeoxCer exhibited pronounced accumulation in the VAT of obese T2D individuals as compared to non-T2D obese subjects ^9^. We did measure DeoxCer lipids in the cohort 2 samples, where they were not identified among the DALs between morbid obese and lean individuals (Figure 4a), further promoting the concept that these lipids are predominantly associated with T2D rather than with obesity alone. Further investigations will be required to explore whether DeoxCer lipid content of adipose tissue will be altered by the weight loss following GBS.

Importantly, in this study we established the lipid signatures for the two types of white adipose tissue: subcutaneous, analysed in cohort 1, and visceral in the cohort 2. Recent comparisons of SAT and VAT lipid content suggested depot-specific lipid features ^9, 11, 24^. In agreement with these reports, we observed higher levels of Cer in VAT in comparison to SAT of both lean and obese origin (Figures 2c and 4b). Moreover, our data reveal elevated levels of CL in SAT in comparison to VAT (Figures 2c and 4b), suggesting a more dynamic mitochondrial activity in SAT. Taken together, our data suggest a strong perturbation of the sphingolipid metabolism in morbid obese patients that is partly counteracted by a GBS intervention.

It should be noted that one drawback of our cohort 1 lipidomic analyses of SAT biopsies is a relatively low number of samples (n=5 of paired SAT samples, total of 10), stemming from the invasive character of the SAT sampling post-GBS. Although the longitudinal design of the study allowing for paired comparisons of the samples derived from the same patients compensated, to some extent, this cohort size limitation, it still impinges on the statistical significance of the obtained results. In conclusion, our results demonstrate a massive rearrangement of the human serum and SAT lipidome in morbid obese individual. Strikingly, we report an improved sphingolipid signature along with a major overhaul of the most abundant glycerophospholipids (PC and PI) following GBS. The comparison of the serum and SAT lipid landscape highlights tissue-specific lipid modifications, although the follow-up studies on a higher number of subjects will be required to confirm this observation. Further examinations are warranted to determine the mechanistic link between long-term clinical benefits associated with surgically induced weight loss and the reported changes of lipid homeostasis.

## Materials and Methods

### Study design, subject characteristics and sample collections

#### Human cohort 1

The clinical characteristics of the individuals included in the cohort 1 have been previously reported by us in ^14^. Samples were obtained from participants with written informed consent. The study was conducted according to the ethical principles for medical research involving human subjects released by the Declaration of Helsinki and had ethics local committee approval. In this work we only included the participants with morbid obesity (defined as BMI ≥ to 40 kg.m^-2^) pre- and post-GBS (see Table supplement 1). The enrolled subjects were non-smokers and exhibited no arterial hypertension (blood pressure < 140/90 mmHg) or diabetes mellitus (HbA_1c_ ≤ 5.8% (40 mmol/mol)). All participants underwent an initial screening visit that comprised a physical examination, blood pressure measurements, euglycemic-hyperinsulinemic clamp test, routine blood chemistry, and serum sampling between 8 and 9 AM following over-night fast. SAT biopsies were taken during planned Roux-en-Y Gastric Bypass with a gastric pouch of 20-30 cm^3^ and an alimentary limb of 150 cm and a biliopancreatic limb of 75 cm. One-year post-GBS, all the participants were subjected again to a final physical examination, euglycemic-hyperinsulinemic clamp test, SAT biopsy, routine blood chemistry, and serum sampling at 8-9 AM following overnight fasting.

#### Human cohort 2

The clinical characteristics of the individuals included in the cohort 2 have been reported by us in ^9^. Samples were obtained from all participants with written informed consent. The study conformed to the Declaration of Helsinki and the experimental protocol (‘DIOMEDE’) was approved by the Ethical Committee SUD EST IV (Agreement 12/111) and performed according to the French legislation (Huriet’s law). In this work, we only analysed the serum and VAT samples from lean control and morbid obese non-diabetic donors who had HbA1c levels inferior to 48 mmol/mol, fasting glycemia inferior to 7 mmol/L, and were not diagnosed with type 2 diabetes (T2D) (see Table supplement 2). VAT biopsies from the same participants were taken during planned bariatric surgery. Serum samples were collected between 8 and 10 AM, following over-night fasting.

### Serum and white adipose tissue sample preparation

Blood samples were collected in clot-activator vacutainers and immediately processed for routine blood chemistry reported in Tables supplement 1 and 2. Serum was prepared by blood centrifugation (10 min, 1650 x g, 4 °C) and stored at −80 °C until lipid extraction and analyses. White adipose tissue biopsies (SAT or VAT) were stored at −80 °C until lipid extraction and analysis.

### Lipidomic experiments

#### Materials for lipid extraction

Synthetic lipid standards [PC 12:0/12:0 (850335), PE 17:0/14:1 (LM-1104), PI 17:0/14:1 (LM-1504), PS 17:0/14:1 (LM-1304), Cer d18:1/17:0 (860517), SM d18:1/12:0 (860583), HexCer d18:1/8:0 (860540)] were from Avanti Polar Lipids Inc. MTBE (methyl-tert-butyl ether) and methylamine (33% in absolute ethanol) were purchased from Sigma Aldrich. Chloroform, methanol, n-butanol and ammonium molybdate were from Acros Organics. LC-MS grade methanol, water and ammonium acetate were from Fluka. HPLC-grade chloroform was purchased from Acros Organics. Monopotassium phosphate, L-ascorbic acid, 70% perchloric acid, hexane, methyl acetate and acetonitrile were from Merck.

#### Serum lipid extraction procedure

Lipid extracts were prepared using a modified MTBE extraction protocol with addition of internal lipid standards ^9, 25^. Briefly, 100 µL serum was used, 360 µL methanol and a mix of internal standards were added (400 pmol PC 12:0/12:0, 1000 pmol PE 17:0/14:1, 1000 pmol PI 17:0/14:1, 3300 pmol PS 17:0/14:1, 2500 pmol SM d18:1/12:0, 500 pmol Cer d18:1/17:0 and 100 pmol HexCer d18:1/8:0). After addition of 1.2 mL of MTBE (methyl tert-butyl ether), samples were placed for 10 minutes on a multitube vortexer at 4°C followed by incubation for 1 hour at room temperature (RT) on a shaker. Phase separation was induced by addition of 200 µL MS-grade water. After 10 minutes at RT, samples were centrifuged at 1000 *g* for 10 minutes. The upper (organic) phase was transferred into a 13 mm glass tube and the lower phase was re-extracted with 400 µL artificial upper phase [MTBE/methanol/H_2_O (10:3:1.5, v/v/v)]. The combined organic phases were dried in a vacuum concentrator (CentriVap, Labconco). Lipid extracts derived from MTBE extraction were resuspended in 750 µL chloroform: methanol (1:1), sonicated and vortexed. Resuspended lipids were divided in 3 aliquots. One aliquot was used for glycerophospholipid analysis, a second one for phosphorus assay, and the third aliquot was treated by mild alkaline hydrolysis to enrich for sphingolipids, according to the method by Clarke ^26^. Briefly, 1 mL freshly prepared monomethylamine reagent [methylamine/H_2_O/n-butanol/methanol (5:3:1:4, (v/v/v/v)] was added to the dried lipid extract and then incubated at 53°C for 1 hour in a water bath. Lipids were cooled to RT and then dried. For desalting, the dried lipid extract was resuspended in 300 µL water-saturated n-butanol and then extracted with 150 µL H_2_O. The organic phase was collected, and the aqueous phase was re-extracted twice with 300 µL water-saturated n-butanol. The organic phases were pooled and dried in a vacuum concentrator.

#### SAT lipid extraction procedure

Lipid extracts were prepared using a modified protocol of the 3-phase liquid extraction method ^27^ with addition of internal lipid standards. Briefly, 30 mg of tissue were homogenized in N_2_-cold condition (Precellys24 Bertin Instruments) in presence of zirconium oxide beads CK14 (Labgene Scientific) and 200 µL methanol:dichloromethane solution (1:2). After addition of 1 mL of hexane, 1 mL of methyl acetate and 0.75 mL of acetonitrile, samples were vortexed at room temperature and centrifuged at 2000 g for 5 min, resulting in the separation of three distinct phases. The upper organic phase was collected, and neutral lipid extracts were dried in a vacuum concentrator. The middle layer was re-extracted with 1 mL of hexane and the bottom phase, containing the polar lipids, was collected and dried in a vacuum concentrator. Polar lipid extracts were resuspended in 550 µL chloroforme:methanol (1:1) and divided in 2 aliquots. One aliquot was used for phosphorus assay, and the second one was treated by mild alkaline hydrolysis to enrich for sphingolipids as described above.

#### Determination of total phosphorus

One hundred µL of the total lipid extract, resuspended in chloroform/methanol (1:1), were placed into 13 mm disposable pyrex tubes and dried in a vacuum concentrator. Zero, 2, 5, 10, 20 µL of a 3 mmol/L KH_2_PO_4_ standard solution were placed into separate pyrex tubes. To each tube, distilled water was added to reach 20 µL of aqueous solution. After addition of 140 µL 70% perchloric acid, samples were heated at 180°C for 1 hour in a chemical hood. Then, 800 µL of a freshly prepared solution of water, ammonium molybdate (100 mg/8 mL H_2_O) and ascorbic acid (100 mg/6 mL H_2_O) in a ratio of 5:2:1 (v/v/v) were added. Tubes were heated at 100°C for 5 minutes with a marble on each tube to prevent evaporation. Tubes were cooled at RT for 5 minutes. One hundred µL of each sample was then transferred into a 96-well microplate and the absorbance at 820 nm was measured.

#### Phospho- and sphingolipid analysis by mass spectrometry

Mass spectrometry analysis for the quantification of phospho- and sphingolipid species was performed using multiple reaction monitoring on a TSQ Vantage Extended Mass Range Mass Spectrometer (ThermoFisher Scientific), equipped with a robotic nanoflow ion source (Triversa Nanomate, Advion Biosciences) as previously described ^9^. Optimized fragmentation was generated using appropriate collision energies and s-lens values for each lipid class. Mass spectrometry data were acquired with TSQ Tune 2.6 SP1 and treated with Xcalibur 4.0 QF2 software (ThermoFisher Scientific). Lipid quantification was carried out using an analysis platform for lipidomics data hosted at EPFL Lausanne Switzerland http://lipidomes.epfl.ch/. Quantification procedure was described in ^28^. Dried lipid extracts were resuspended in 250 µL MS-grade chloroform/methanol (1:1) and further diluted in either chloroform/methanol (1:2) plus 5 mmol/L ammonium acetate (negative ion mode) or in chloroform/methanol/H_2_O (2:7:1) plus 5 mmol/L ammonium acetate (positive ion mode).

#### Data quantification and statistical analyses

Lipid concentrations were calculated relative to the relevant internal standards and normalized to the total phosphate content of each total lipid extract. To compare between different lipid samples, relative lipid concentrations were normalized to the total lipid content of each lipid extract (mol%). Additional data processing (filtering, normalization, transformation, scaling), statistical analyses, and data plotting were performed using MetaboAnalyst 5.0. ^29^ and Prism Graph Pad 8.0. Statistical tests used for comparison between the groups are indicated in the figure legends. Differences were considered significant for p ≤ 0.05 (*), p ≤ 0.01 (**) and p ≤ 0.001 (***). To determine the clustering, k-NN (nearest neighbours with k clusters) was applied for k = 1, 2, and 3 clusters.

## Data availability

All data generated or analysed during this study are included in this published article (and its supplementary information files).

## Contribution statement

FS performed lipidomics experiments; FS and SC analyzed the lipidomic data and performed statistical analyses; MCBM and JPM assisted with the lipidomic studies; ZP conceptualized the cohort 1, ZP and MJ provided the samples; EF conceptualized the cohort 2 and provided the samples. GdA and HR contributed to the lipidomics conceptualization. CD designed and coordinated the study; FS and CD drafted the manuscript. All authors contributed to the manuscript preparation and approved the final version.

## Supplementary material

**Figure supplement 1.**
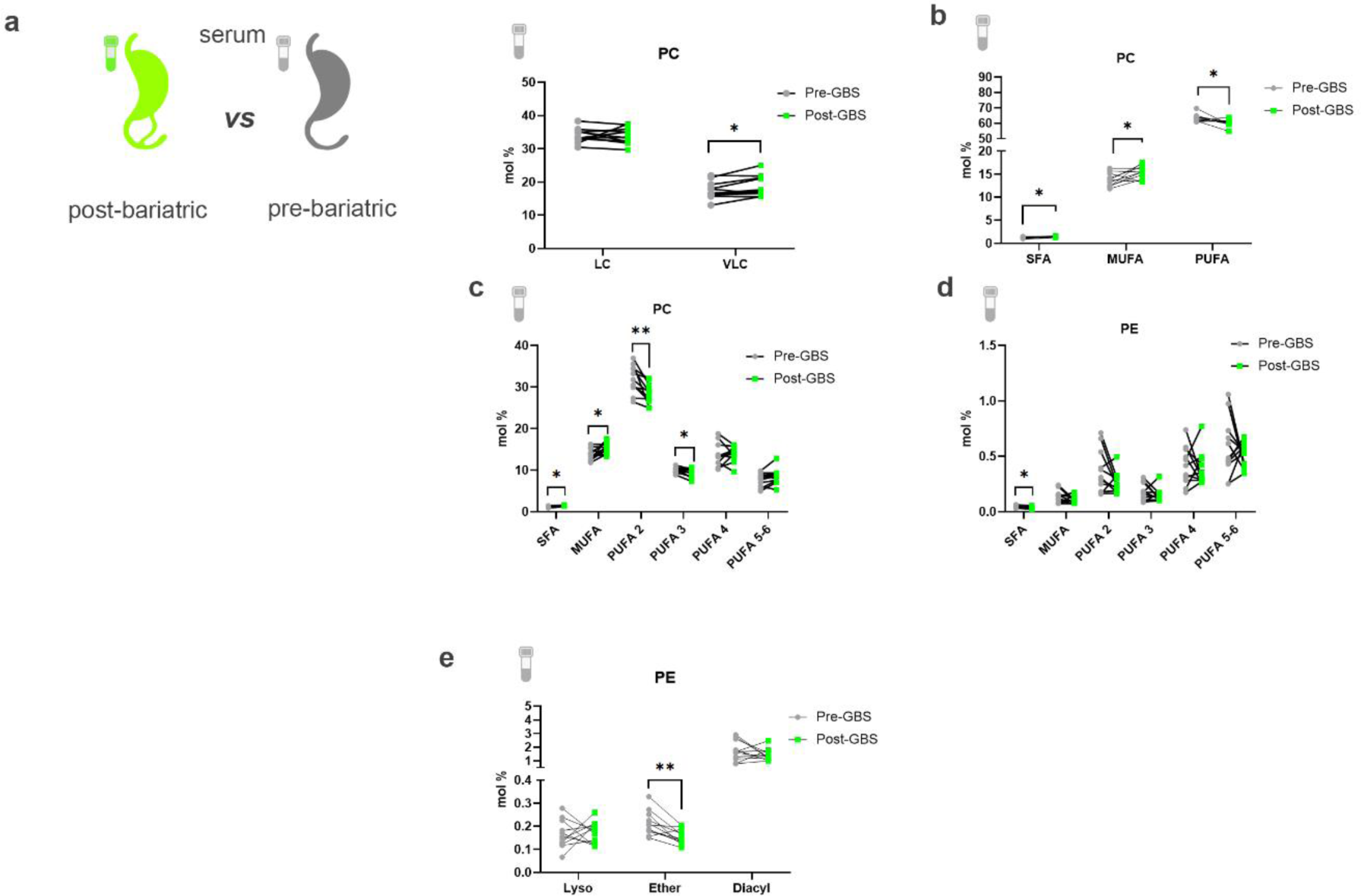
Comparison of differentially abundant serum lipids measured in obese individuals before and after bariatric surgery. **(a)** Relative PC level changes (mol%) in sera collected before and after GBS, clustered according to the chain length. LC (long chain C28-34), VLC (very long chain C38-44). **(b)** Relative PC level changes (mol%) in sera collected before and after GBS, represented according to the class of saturation degree: saturated fatty acids (SFA), monounsaturated fatty acids (MUFA) and polyunsaturated fatty acids (PUFA). **(c-d)** Relative PC **(c)** and PE **(d)** level changes (mol%) in sera collected before and after GBS, clustered according to the degree of saturation. (**e**) Relative PE level changes (mol%) in serum collected before and after GBS, represented according to the nature of the fatty acid linkage (diacyl vs alkyl-acyl (ether) or monoacyl (lyso)). Statistics for (**c-e**) are paired student’s *t*-test. Data for serum (n = 11) pre-GBS and post-GBS are represented as mean ± SEM. * p < 0.05; ** p < 0.01.

**Figure supplement 2.**
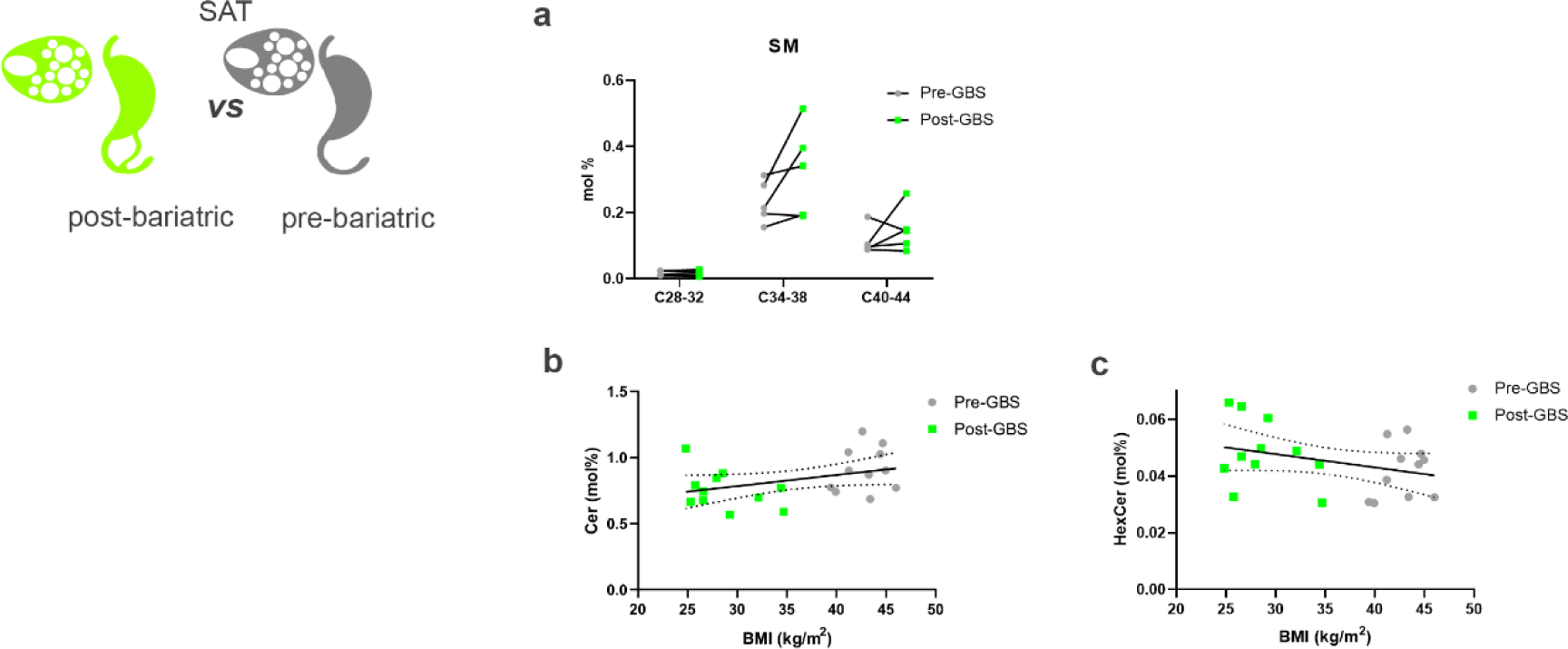
Comparison of differentially abundant SAT lipids measured in obese individuals before and after bariatric surgery. (**a**) Relative SM level changes (mol%) in sera collected before and after GBS (n = 5), represented according to the chain length. Data are represented as mean ± SEM. (**b-c**) Association between the relative levels of ceramides (**b**), or HexCer (**c**) detected in sera, and the BMI of the subjects before and after GBS (n = 22, Spearman correlation, for the ceramides R = 0.334, p = 0.1291, for the HexCer, R = −0.237 p = 0.289).

## Tables supplement

**Table supplement 1.**
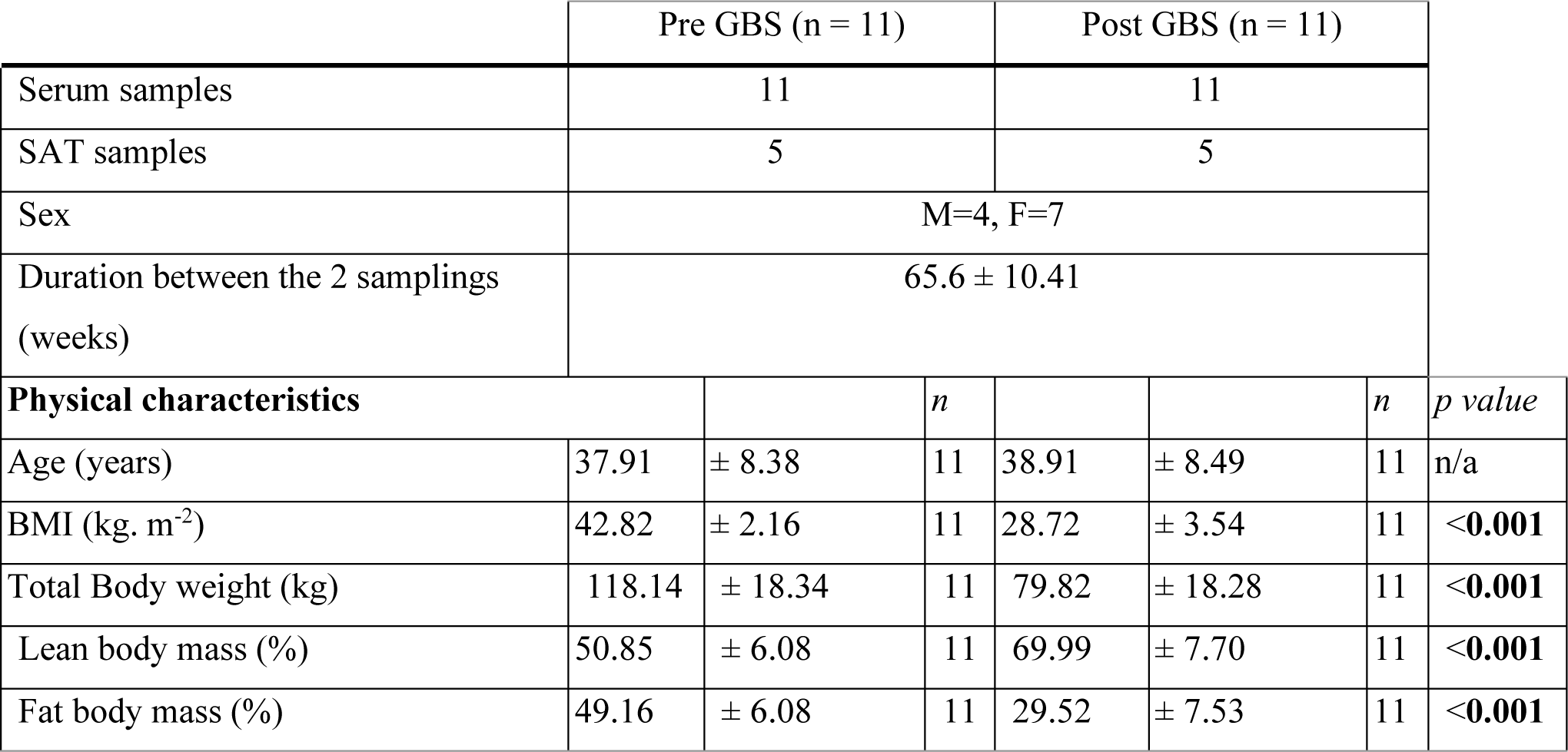
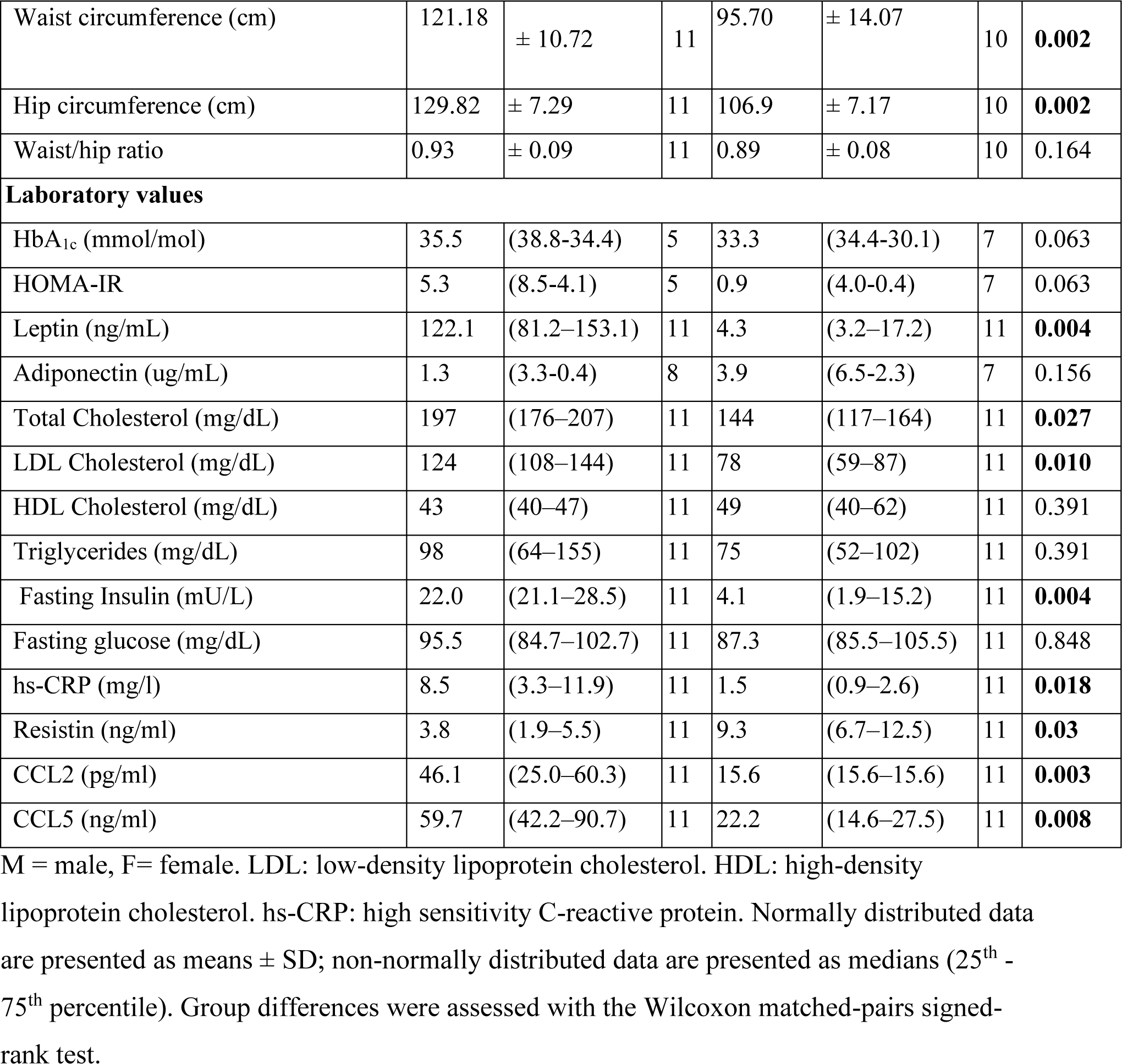
Participant characteristics (cohort 1)

**Table supplement 2.** Participant characteristics (cohort 2)

|  | Serum |  |  |  |  | VAT |  |  |  |  |
| --- | --- | --- | --- | --- | --- | --- | --- | --- | --- | --- |
|  | Control (n = 5) |  | Obese (n = 16) |  | *p value* | Control (n = 7) |  | Obese (n = 11) |  | *P value* |
| Sex (M/F) | 3/2 |  | 6/10 |  |  | 3/4 |  | 4/7 |  |  |
| Age (years) | 52 | (46.50-63.50) | 45 | (37.00-53.20) | 0.1339 | 51 | (40-58) | 45 | (37-55) | 0.675 |
| BMI (kg. m<sup>-2</sup>) | 24.01 | (20.12-24.74) | 45.34 | (42.86-47.24) | **<0.0001** | 22.898 | (18.94-25.10) | 46.65 | (44.06-48.28) | **<0.0001** |
| HbA<sub>1c</sub> (mmol/mol) | ND |  | 38 | (37-38) | n/a | ND |  | ND |  | n/a |
| Fasting glycemia (mmol/L) | ND |  | 5 | (4.7-5.2) | n/a | ND |  | ND |  | n/a |

M = male; W = female; visceral adipose tissue (VAT); not determined (ND); Not applicable: (n/a). Data are presented as medians (25<sup>th</sup> - 75<sup>th</sup> percentile). Group differences were assessed with the Mann-Whitney *U* test.

**Table supplement 3.**
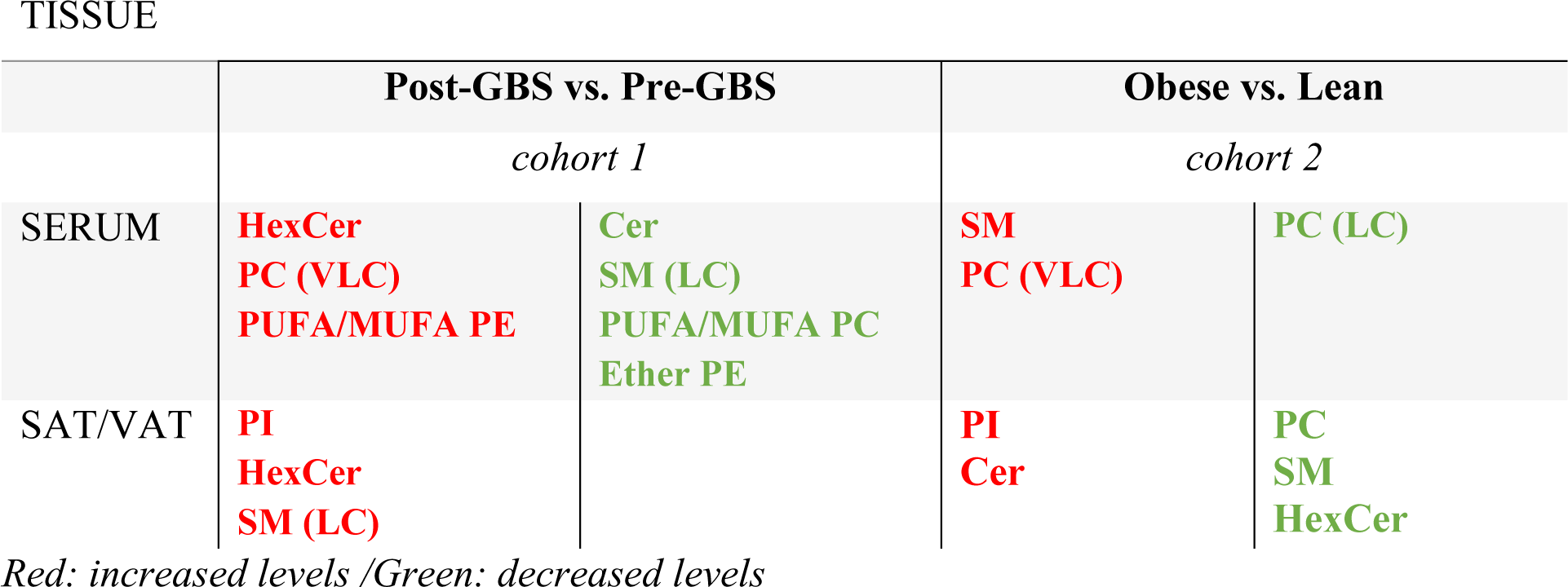
Comparison of the major lipid classes changes in the patient cohorts 1 and 2.

## References

1. Sjöström, L.; Lindroos, A.-K.; Peltonen, M., et al., Lifestyle, Diabetes, and Cardiovascular Risk Factors 10 Years after Bariatric Surgery. New England Journal of Medicine 2004, 351 (26), 2683–2693.

2. Sjöström, L.; Narbro, K.; Sjöström, C. D., et al., Effects of bariatric surgery on mortality in Swedish obese subjects. N Engl J Med 2007, 357 (8), 741–52.

3. Bradley, D.; Magkos, F.; Klein, S., Effects of Bariatric Surgery on Glucose Homeostasis and Type 2 Diabetes. Gastroenterology 2012, 143 (4), 897–912.

4. Lee, G.; Park, Y. S.; Cho, C., et al., Short-term changes in the serum metabolome after laparoscopic sleeve gastrectomy and Roux-en-Y gastric bypass. Metabolomics 2021, 17 (8), 71.

5. Mikhalkova, D.; Holman, S. R.; Jiang, H., et al., Bariatric Surgery-Induced Cardiac and Lipidomic Changes in Obesity-Related Heart Failure with Preserved Ejection Fraction. Obesity (Silver Spring) 2018, 26 (2), 284–290.

6. Kayser, B. D.; Lhomme, M.; Dao, M. C., et al., Serum lipidomics reveals early differential effects of gastric bypass compared with banding on phospholipids and sphingolipids independent of differences in weight loss. Int J Obes (Lond) 2017, 41 (6), 917–925.

7. Arora, T.; Velagapudi, V.; Pournaras, D. J., et al., Roux-en-Y Gastric Bypass Surgery Induces Early Plasma Metabolomic and Lipidomic Alterations in Humans Associated with Diabetes Remission. PLoS One 2015, 10 (5), e0126401.

8. Graessler, J.; Bornstein, T. D.; Goel, D., et al., Lipidomic profiling before and after Roux-en-Y gastric bypass in obese patients with diabetes. Pharmacogenomics J 2014, 14 (3), 201–7.

9. Hannich, J. T.; Loizides-Mangold, U.; Sinturel, F., et al., Ether lipids, sphingolipids and toxic 1-deoxyceramides as hallmarks for lean and obese type 2 diabetic patients. Acta Physiol (Oxf) 2021, 232 (1), e13610.

10. Yin, X.; Willinger, C. M.; Keefe, J., et al., Lipidomic profiling identifies signatures of metabolic risk. EBioMedicine 2020, 51, 102520.

11. Lange, M.; Angelidou, G.; Ni, Z., et al., AdipoAtlas: A reference lipidome for human white adipose tissue. Cell Rep Med 2021, 2 (10), 100407.

12. Sinturel, F.; Spaleniak, W.; Dibner, C., Circadian rhythm of lipid metabolism. Biochem Soc Trans 2022.

13. Petrenko, V.; Sinturel, F.; Riezman, H.; Dibner, C., Lipid metabolism around the body clocks. Prog Lipid Res 2023, 91, 101235.

14. Montecucco, F.; Lenglet, S.; Quercioli, A., et al., Gastric bypass in morbid obese patients is associated with reduction in adipose tissue inflammation via N-oleoylethanolamide (OEA)-mediated pathways. Thromb Haemost 2015, 113 (4), 838–50.

15. Huang, H.; Kasumov, T.; Gatmaitan, P., et al., Gastric bypass surgery reduces plasma ceramide subspecies and improves insulin sensitivity in severely obese patients. Obesity (Silver Spring) 2011, 19 (11), 2235–40.

16. Hanamatsu, H.; Ohnishi, S.; Sakai, S., et al., Altered levels of serum sphingomyelin and ceramide containing distinct acyl chains in young obese adults. Nutr Diabetes 2014, 4 (10), e141.

17. Tulipani, S.; Palau-Rodriguez, M.; Miñarro Alonso, A., et al., Biomarkers of Morbid Obesity and Prediabetes by Metabolomic Profiling of Human Discordant Phenotypes. Clin Chim Acta 2016, 463, 53–61.

18. Lemaitre, R. N.; Yu, C.; Hoofnagle, A., et al., Circulating Sphingolipids, Insulin, HOMA-IR, and HOMA-B: The Strong Heart Family Study. Diabetes 2018, 67 (8), 1663–1672.

19. Dashti, M.; Kulik, W.; Hoek, F., et al., A phospholipidomic analysis of all defined human plasma lipoproteins. Sci Rep 2011, 1, 139.

20. Aburasayn, H.; Al Batran, R.; Ussher, J. R., Targeting ceramide metabolism in obesity. Am J Physiol Endocrinol Metab 2016, 311 (2), E423–35.

21. Raichur, S.; Brunner, B.; Bielohuby, M., et al., The role of C16:0 ceramide in the development of obesity and type 2 diabetes: CerS6 inhibition as a novel therapeutic approach. Mol Metab 2019, 21, 36–50.

22. Juchnicka, I.; Kuźmicki, M.; Szamatowicz, J., Ceramides and Sphingosino-1-Phosphate in Obesity. Frontiers in Endocrinology 2021, 12.

23. Sokolowska, E.; Blachnio-Zabielska, A., The Role of Ceramides in Insulin Resistance. Front Endocrinol (Lausanne) 2019, 10, 577.

24. Zacharia, A.; Saidemberg, D.; Mannully, C. T., et al., Distinct infrastructure of lipid networks in visceral and subcutaneous adipose tissues in overweight humans. Am J Clin Nutr 2020, 112 (4), 979–990.

25. Matyash, V.; Liebisch, G.; Kurzchalia, T. V., et al., Lipid extraction by methyl-tert-butyl ether for high-throughput lipidomics. J Lipid Res 2008, 49 (5), 1137–46.

26. Clarke, N. G.; Dawson, R. M., Alkaline O leads to N-transacylation. A new method for the quantitative deacylation of phospholipids. Biochem J 1981, 195 (1), 301–6.

27. Vale, G.; Martin, S. A.; Mitsche, M. A., et al., Three-phase liquid extraction: a simple and fast method for lipidomic workflows. J Lipid Res 2019, 60 (3), 694–706.

28. Pietiläinen, K. H.; Sysi-Aho, M.; Rissanen, A., et al., Acquired obesity is associated with changes in the serum lipidomic profile independent of genetic effects--a monozygotic twin study. PLoS One 2007, 2 (2), e218.

29. Pang, Z.; Chong, J.; Zhou, G., et al., MetaboAnalyst 5.0: narrowing the gap between raw spectra and functional insights. Nucleic Acids Res 2021, 49 (W1), W388–w396.

